# Measuring Multi-Joint Upper Limb Proprioceptive Position Sense in Children with Unilateral Spastic Cerebral Palsy using 3D Kinematics

**DOI:** 10.1101/2024.09.30.24314648

**Authors:** Shivakeshavan Ratnadurai-Giridharan, Dalina Delfing, Maxime T. Robert, Tomoko Kitago, Andrew M. Gordon, Kathleen M. Friel

## Abstract

**Background:** Proprioceptive position sense (PPS), which provides the awareness of the positions of one’s limbs in space, is compromised in children with unilateral spastic cerebral palsy (USCP). Relationships between decreased PPS, skill acquisition, and performance are not well understood partly due to differences between measurement modalities of PPS. During assessments, individual joints are often constrained to a single plane of movement. We introduce ways to estimate PPS, simultaneously from multiple upper limb joints during unconstrained pose matching tasks, using 3D kinematics.

**Methods:** Nineteen children with USCP, 14 age-matched typically developing children (TDC) and, 16 typically developed adults (TDA) were recruited. Participants performed contralateral upper limb pose matching tasks while blindfolded. Using 3D kinematic data extracted by a VICON Nexus motion-capture system, three ways of measuring matching performance were calculated: joint angle symmetry, joint distance symmetry, and orientation symmetry. Mann-Whitney U tests were used to statistically compare scores between groups. Spearman’s rank correlation was used to investigate relationships between PPS symmetry measures and hand function in children with USCP.

**Results:** Children with USCP presented significantly lower angle, distance, and orientation symmetries compared to TDC. TDA scores were not statistically significantly different compared to TDC. In children with USCP, orientation symmetry measures strongly correlated with several clinical hand function scores such as the box and blocks test.

**Conclusions:** We introduced a new way to conveniently measure multi-joint PPS. Children with USCP presented lower PPS estimates compared to TDC. We also observed that orientation symmetry (limb co-ordination) strongly correlates with hand function.

## 1 Introduction

Proprioceptive position sense (PPS) is the awareness of the positional information regarding the body and limbs relative to each other in space. It is a critical component of the sensorimotor system that enables the development of motor skills [1]. In neurological disorders affecting movement, such as unilateral spastic cerebral palsy (USCP), proprioceptive abilities of the upper extremities (UE) including PPS are known to be affected leading to reduced balance and coordination [2, 3]. Studies have shown that proprioception can be trained and improved to a certain extent [4]. However, the relationship between proprioception and skilled movements is still not clearly understood [4–6]. Studies examining the relationship between proprioception and hand function in affected populations, have reported ranges varying from poor to high correlation between proprioception and hand function [7–10]. This discrepancy may have been in part due to the differences in how PPS was measured in these studies [11].

Rather than a purely sensory modality, proprioceptive position sense (PPS) is a complex process that involves the integration of both afferent and efferent activity [12]. Sensory information from muscle spindles, tendons, joints, and skin along with motor neuron activity are combined to provide a predictive estimation of a limb’s or body’s pose. The complexity of this can be further appreciated by considering that in the real world, multiple joints need to be coordinated for proper function. This requires accurate predictions of the position and pose of body parts by integrating afferent and efferent information across multiple joints simultaneously. However most of the current methods for measuring PPS primarily consider a single joint at a time.

To better understand the role of PPS in upper limb functional movement and its development, there needs to be a comprehensive way to measure it across multiple joints simultaneously. There have been many different approaches that have attempted to measure PPS but, typically the focus is on a single joint. Common approaches can generally be classified into three categories [12, 13] 1) Movement threshold detection: where the limb is passively moved in small increments and the participant is asked to detect when the arm has actually been displaced [14, 15]. This is typically done using a manipulandum apparatus or goniometer. It requires many trials that may take hours for a reliable estimation of PPS; 2) Active or passive joint position reproduction assessments: where a participant is asked to match or detect a reference target pose by actively moving their arm or having it passively moved for them [16, 17]. The outcome of this assessment is a single joint angle which is measured manually using a goniometer or through sensors on a robotic arm. This approach typically requires fewer trials than the other approaches; 3) Passive direction discrimination: where the participant’s arm is passively moved and the participant is asked to state the perceived direction of movement [18, 19]. This assessment also requires a specialized apparatus and multiple trials to get a good representation of estimated PPS. The measurement of PPS often requires multiple trials that can take up to a few hours [13]. This becomes a feasibility issue when attempting to measure PPS in children with USCP, who are more prone to fatigue and challenges in staying engaged during assessments [20]. This issue is further compounded when multiple body joints are being assessed. The use of robotics may help in addressing the issue of fatigue by providing body support during PPS estimation. However, a number of studies have suggested that constraining movements during assessments, as often done with robotics, may not be truly representative of natural movement patterns or abilities of the participant [21, 22].

For estimating PPS of the upper extremities in children with USCP, an efficient and reliable method that can estimate the overall PPS of multiple unconstrained joints simultaneously is highly desirable. In order to do this, we model the contralateral joint position reproduction method as a proprioception-centric closed loop feedback system [23,24] as illustrated in Figure 1. In this task the participant is blindfolded and one of their arms are physically moved by the assessor to assume a fixed pose which serves as the reference pose. As seen on the left side of Figure 1, the corresponding PPS (passive) is generated primarily from sensory stimuli. Upon providing a cue, the participant then attempts to match(mirror) the reference pose using their other arm. In this case, active PPS is formed from both sensory information as well as from information pertaining to muscle actuation as modeled in the right of Figure 1. The differences between the perceived poses of the two arms are then cognitively computed and this error provides feedback for correcting the matching arm pose via further muscle actuation. This model is based on a representation of currently known factors that influence PPS [12, 13, 23, 24] associated with joint position reproduction tasks. In this model, we assume that when the perceived error approaches a minimum, the participant ceases further attempts to correct the position of the matching arm. At this point in time quantified comparisons between the reference and the matching arm pose are assumed to be directly proportional to PPS accuracy. With this model in mind, we seek to estimate PPS by comparing the symmetry across contralateral arm poses. We used 3D kinematics captured from an opto-electronic motion capture device to estimate the degree of matching of joint positions during a UE contralateral pose matching task [17]. This way simultaneous measurements are taken across multiple joints when estimating overall PPS ability. Along with joint angle measurements, we introduce two additional metrics to measure the similarity between the contralateral arm poses: joint distance symmetry and joint orientation symmetry respectively. For a given pose, the joint distance symmetry measures the similarity in distance between corresponding joints in the left and right arm against a common reference point such as the neck. The orientation symmetry measures the alignment between the left and right arms during the pose matching task, i.e. the degree to which the left and right UE align along the same plane. These additional metrics are needed because in 3D space a single metric is insufficient to capture the similarity of two arm poses that are mirrored. This is discussed in further detail in Section 2.4. For this study, we acquired PPS measures from three different populations, children with USCP, typically developing children (TDC) and, typically developed adults (TDA). We also investigated how these measured metrics of PPS correlated with various measures of hand function in children with USCP.

**Figure 1:**
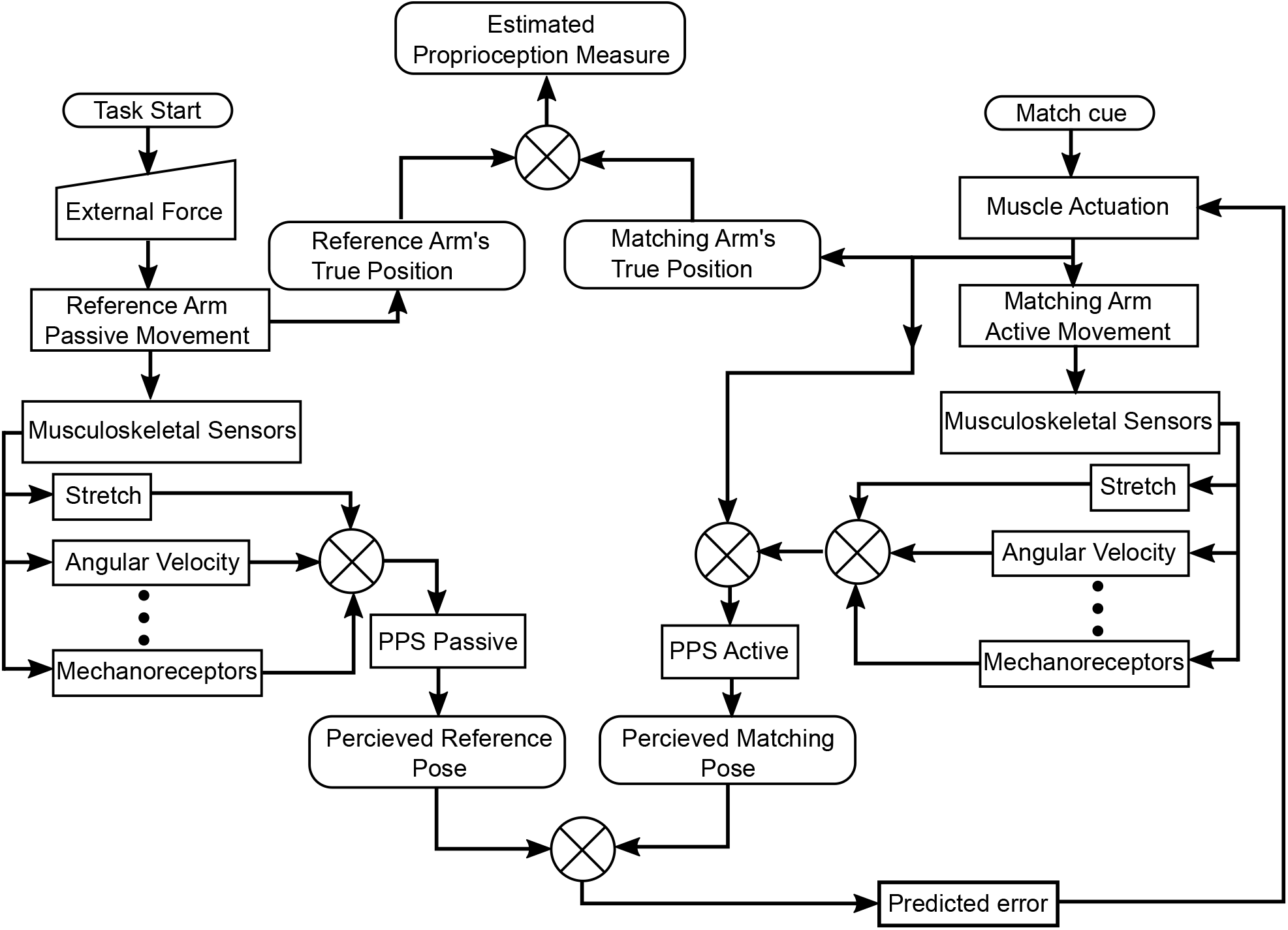
A schematic of the closed loop process involved in proprioceptive position sense estimation for the contralateral joint/pose position reproduction task.

## 2 Methods

### 2.1 Study Population

We recruited a convenience sample of 19 children with unilateral spastic cerebral palsy (11 ± 3.6 years) from a group of participants in an upper limb rehabilitation trial. Additionally, we independently recruited 14 typically developing children (TDC) as age matched controls (mean ages of 11 ± 4 years) and 16 adult controls (32 ± 6.55 years) for age based comparisons. Participant demographics are summarized in Table 1.

**Table 1:** Demographic break up of study participants by groups, age, gender and race.

| Group | Sample size | Age<br>(yrs) | Gender<br>(% Female) | Race(%) |  |  |  |
| --- | --- | --- | --- | --- | --- | --- | --- |
|  |  |  |  | Asian | Black | White | Other |
| USCP | 19 | $11 \pm 3.6$ | 30 | 20 | 20 | 50 | 10 |
| TDC | 14 | $11 \pm 4$ | 60 | 30 | 0 | 50 | 20 |
| TDA | 16 | $32 \pm 6.25$ | 83 | 39 | 5.5 | 50 | 5.5 |

Consent was obtained from all participants or their guardians. Additionally, children under eighteen years of age provided their assent to participate in the study.

### 2.2 Proprioceptive Position sense assessment task

Participants were asked to sit comfortably upright on a chair and were blindfolded. One of the upper limbs was guided to a reference position as close as possible to the illustrated poses (Figure 2), while minimizing discomfort. Regardless of the exact pose assumed by a participant, we included data from all participants, as our primary goal was to measure similarities between the reference limb pose and the matching limb’s pose. Participants were asked to hold their current pose and then match (mirror) it with their other arm. Children with USCP were tested twice for each of the two poses. First: while matching the pose with their less affected hand when the affected arm’s pose was the reference. Second: with their affected hand when the less affected arm’s pose was the reference. Typically developing children were asked to perform the matching only with their preferred hand. All participants were requested to maintain the matched poses for a period of 5 seconds. Participants were verbally cued to perform the tasks with the commands “Match”, “Hold” and “Relax”, respectively. The two poses shown in Figure 2 were picked from other poses tested during a pilot study. They were selected on the assumption of the ”Muscles” flexing pose being more natural and commonly occuring, while the ”Powerbars” lifting pose was observed to be more challenging to assume and was less commonly used day-to-day. This allows us to determine if differences in poses to be assumed during matching affect our estimation of PPS.

**Figure 2:**
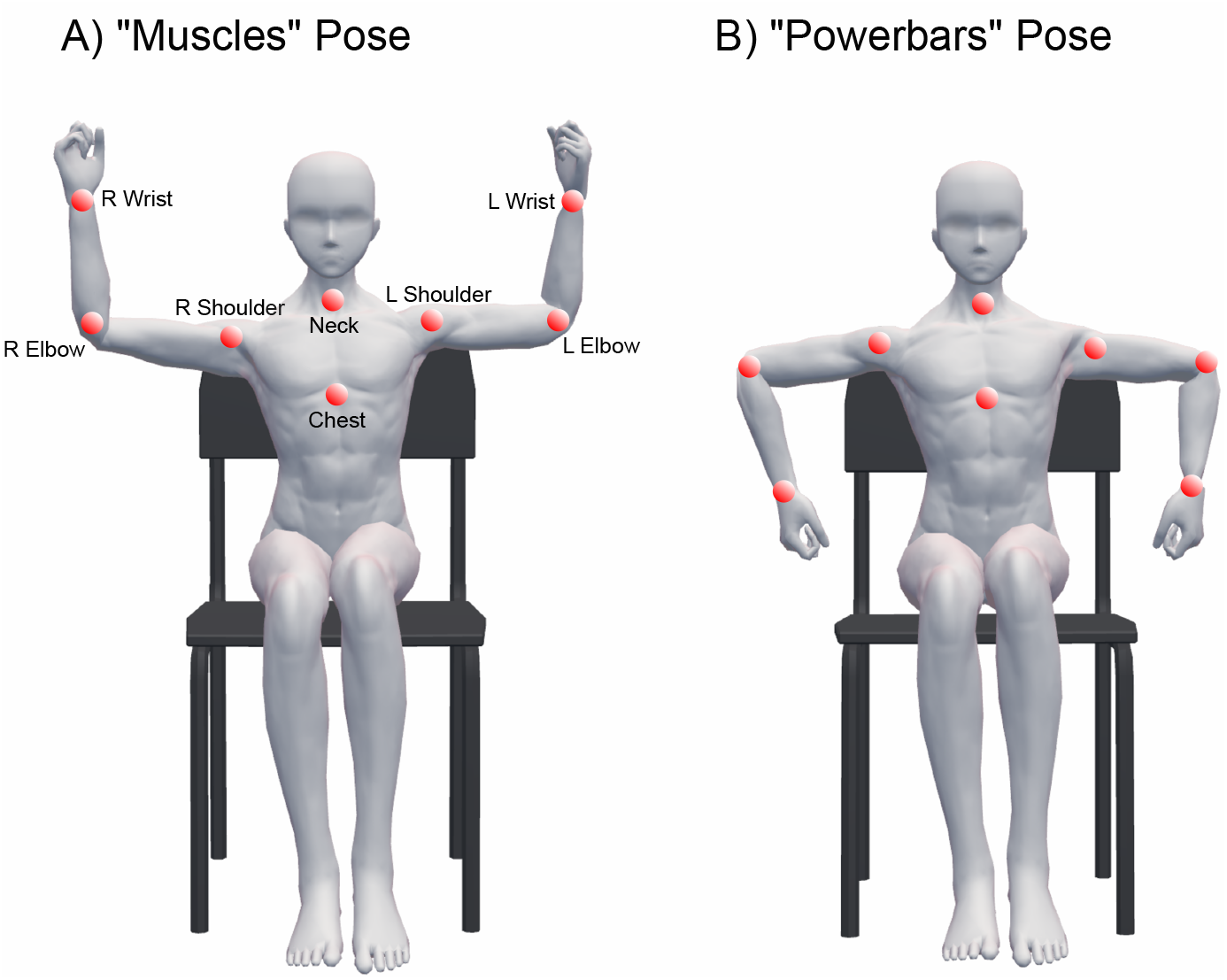
The matching poses to be assumed with tracked joints indicated by the red balls: **A)** A natural ”Muscles” flex pose and **B)** A less natural ”Powerbars” lifting pose

### 2.3 Kinematic Data

Three dimensional kinematic data was collected from participants using an optoelectronic 10-camera 3D motion capture system (VICON Nexus v 2.2.1, Lake Forest, CA). One centimeter diameter reflective markers were placed at twelve of the following anatomical landmarks: the neck, mid-sternum, shoulders (acromion), medial and lateral epicondyles of the elbow and, the ulnar and radial styloid process of the wrists. Kinematic data was captured at a sampling rate of 100 Hz.

For analysis, 3D kinematic data was low-pass filtered using a 5th order Butterworth with a cutoff frequency of 3 Hz. Occasionally, markers on the elbow were missing for brief periods due to self-occlusion. These missing data points were estimated and restored using cubic spline interpolation. For the analysis of kinematic joint data, elbow positions were calculated as the average position of the lateral and medial epicondyles, while wrist positions were calculated as the average position of the ulnar and radial styloid process. This results in the kinematic tracking of the 8 anatomical locations as specified in Figure 2A.

### 2.4 Proprioceptive Position sense outcome measures

In our study, to measure multi-joint proprioception in 3D space, we asked participant to reproduce the mirrored pose of one arm using the other as shown in Figure 2. In 3D space, the mirroring of the pose is assumed to be the sagittal plane of the participant. It is then possible to project the virtual reflection of one arm across the sagittal plane and check how closely the matching arm is to the virtually reflected arm using a distance measure. However, depending on the assumed trunk posture of the participant, it can be quite challenging to identify this sagittal plane accurately. Using an inaccurate version of the sagittal plane can lead to misleading measures of mirror symmetry. To avoid this issue, we instead use a set of three independent metrics to comprehensively measure aspects of similarity between the two arm poses: angle symmetry, distance symmetry, and orientation symmetry. While the angle and distance symmetries are primarily driven by muscle stretching [25], orientation symmetry primarily measures positional alignment of the UE. As such, this measure is also sensitive to joint rotations. Together, these three independent metrics ensure that similarities between two separate multi-joint upper limb poses can be reliably compared in 3D space. Proof that these symmetry metrics are independent is explored in more detail in Section 3.1.

#### 2.4.1 Distance Symmetry

In the domain of 3D multi-joint pose reproduction, the similarities between joint angles alone are insufficient for determining if two contralateral arm poses are alike. To correctly assess the similarity between two arm poses in a 3D bilateral multi joint pose reproduction task, additional measurements are needed. One other dimension of measuring joint position reproduction is the relative distance of UE joints from a common reference point along the central axis of the body such as the center of the chest or the neck (See Figure 3B). We use the following equation for calculating the distance symmetry for a set of bilateral arm poses.

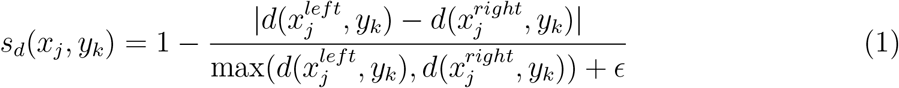

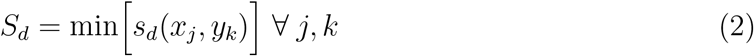

**Figure 3:**
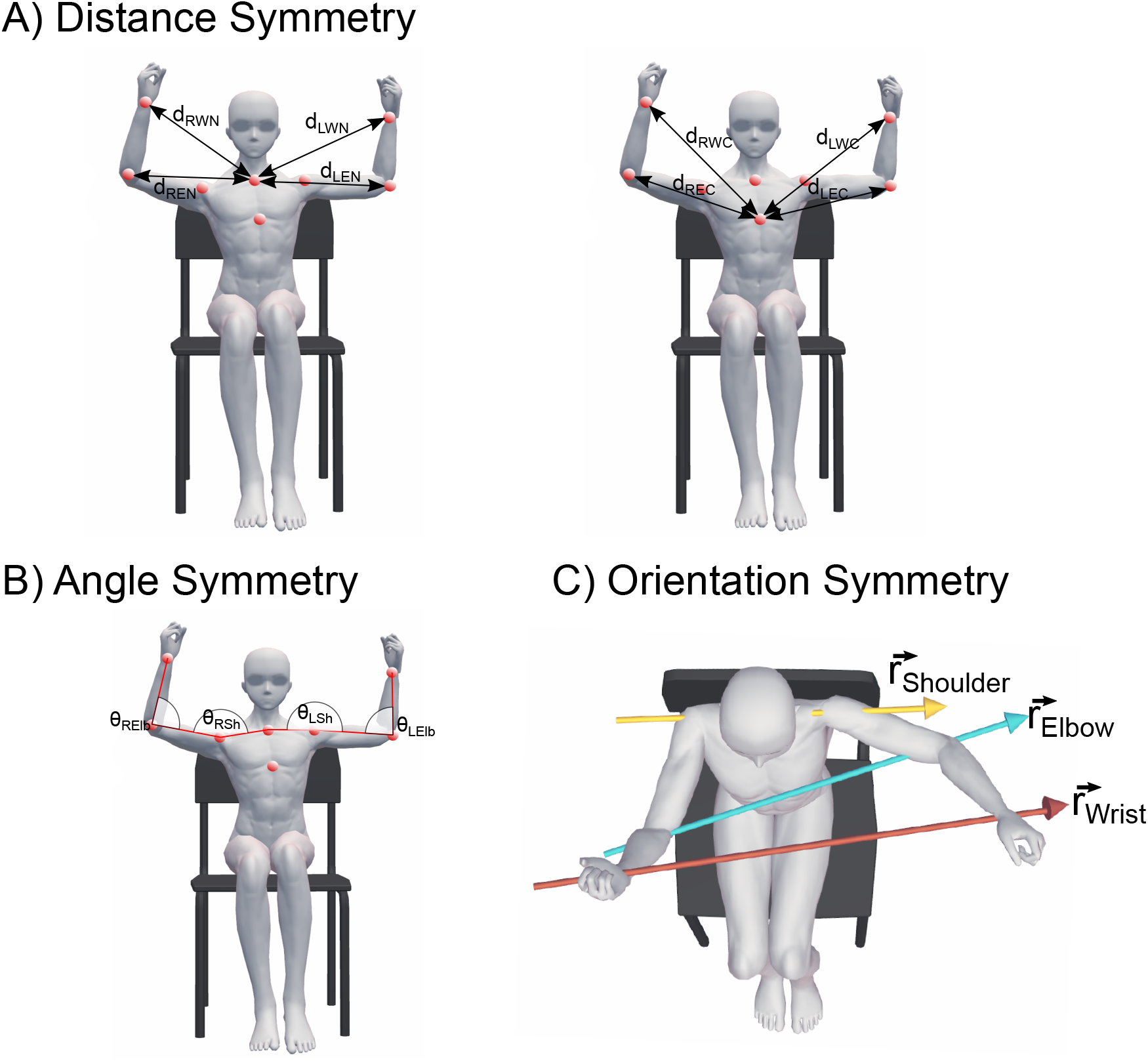
Symmetry measures for estimating proprioception: **A)** Distance symmetry is computed using the corresponding distances of the respective left and right joints from the neck and chest as common reference points. **B)** Angles subtended at the shoulders and elbows are compared between the left and right sides to estimate angle symmetry. **C)** Orientation symmetry measures how aligned multiple joints between the left and right arm are. Symmetric alignment would result in the shoulder, elbow and wrist vectors being more in parallel with one another.

Where, s_*d*_(x_*j*_, y_*k*_) is the distance symmetry between joint j at position x_*j*_ and reference point k at position y_*k*_. S_*d*_ is the overall distance symmetry for the observed pose as a whole.

### 2.4.2 Angle Symmetry

Joint angles of the shoulders and elbows were measured between the reference arm pose (left) and the matching arm pose (right) as shown in Figure 3C. The angles of corresponding joints were then compared pairwise between the two arms as a ratio, where a value of 1 indicated that the angles of a corresponding joint was the same on both sides. The angle symmetry component of PPS was then defined as the minimum of the ratio values calculated for the shoulder and elbow joints. This is explicitly calculated as:

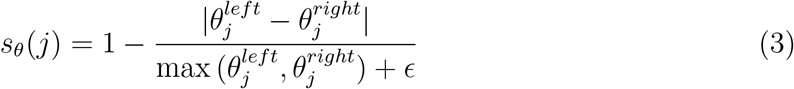

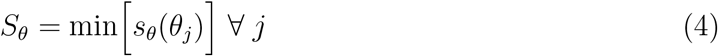

Here, *θ*_*j*_ is the measured angle subtended by joint *j* (shoulder, elbow joints), *s*_*θ*_(_*j*_) is the angle symmetry calculated for joint *j*, and *S*_*θ*_ is the overall angle symmetry for an observed UE pose.

#### 2.4.3 Orientation Symmetry

We introduce one more symmetry metric for measuring similarity of poses to capture the alignment of UE limbs. The orientation symmetry measures the angle differences between a set of vectors as shown in Figure 3D. This metric is needed to complement angle and distance symmetries because, even if distance and angle symmetries are high as seen in Figure 3C, the UE joints of the left and right arms are not oriented or aligned along the same direction. This indicates that the two arms are not matching mirror poses of one another. To measure the degree of alignment of UE joints we calculate the orientation symmetry *S*_*o*_ as:

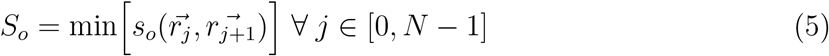

Where,

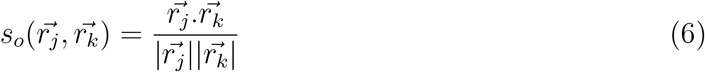

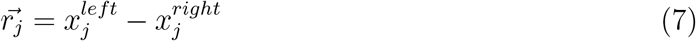

The angle between any two vectors 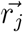 and 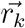 is represented by 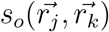. Where, 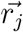 is the vector that connects the left position 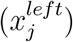 of joint *j* to the corresponding right position 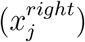 of that respective joint.

### 2.5 Clinical Measurements of Hand function

To investigate the relationship between PPS outcome measures and hand function in children with USCP, we considered the following clinical measurements of hand function.

#### 2.5.1 The Box and Blocks Test

The box and blocks test (BBT) is a gross unimanual dexterity assessment [26]. The BBT apparatus consists of a rectangular box separated into 2 square compartments by a vertical partition. One hundred and fifty colored wooden blocks are placed in one of the compartments. The task involves moving as many blocks as possible with one hand, over the partition into the other empty compartment one block at a time. The score for that particular hand is taken as the number of blocks correctly transferred within 1 minute by the participant.

#### 2.5.2 Assisting Hand Assessment

The assisting hand assessment or AHA measures the degree to which a child with USCP uses their affected hand together with their less affected hand during semi-structured play lasting about 15 minutes [27]. The participant engages in interactive bimanual play across a range of tasks and games while a video recording of the session is taken and later scored offline by a trained evaluator. The scores can range from 22 (affected hand barely used) to 88 (affected hand used similarly to a typical non-dominant hand).

#### 2.5.3 Jebsen-Taylor Test of Hand Function

The Jebsen-Taylor Test of Hand Function (JTTHF) is a standardized test that measures unimanual fine and gross motor function using simulated activities of daily living [28]. The assessment consists of a set of timed tasks such as writing, moving light and heavy objects, picking up small objects, and turning over paper cards. The score for each task is the time taken to complete the task. The total assessment’s score is the sum of the times across all tasks. The maximum allowed time for completion per task is 120 seconds.

#### 2.5.4 Manual ability Classification System

The Manual ability Classification System (MACS) is used to classify how children with cerebral palsy use their hands to handle objects in daily life [29]. It is in the form of a questionnaire that is typically filled by a parent or guardian who knows the child’s actual performance in daily life.

### 2.6 Statistical Methods

PPS symmetry metrics were computed (see Position sense measurement subsection below) on this kinematic time series data. For each trial the median value across time of the respective metric being calculated was taken as the corresponding symmetry’s representative measure. Planned statistical comparisons of PPS symmetry metrics between groups were done with the Mann-Whitney U test. We used a one-tailed test when comparing between children with USCP and TDC as, based on previous studies, we expected children with USCP to exhibit lower levels of PPS. Post-hoc tests were not used when comparing children with USCP and typically developing children, since all pairwise comparisons of metrics between the two groups were statistically significant and in consistent agreement. We used Spearman’s rank correlation to study the relationship between the PPS symmetry measures and hand function as assessed by BBT, JTTHF, MACS and AHA.

## 3 Results

We measured angle, distance and orientation symmetries in all three groups. For the USCP group, the symmetry measures were estimated twice for each task. Once when the pose matching task was performed with the less affected hand, and again when the pose matching task was performed with the affected hand. In Table 2 we report the median and interquartile range (IQR) values of the symmetry values for each group of participants and for each task. It can be observed that a general trend is that the USCP group exhibits lower values of all three symmetries compared to to the TDC group and the adult group. It can also be noted that typically developing children have consistent, but not statistically significant, lower scores compared to the older adult group.

**Table 2:** Median and interquartile range values of Symmetry metric scores for PPS estimation when pose matching is done in children with USCP using their less affected hand, children with USCP using their affected hand, typically developing children using their preferred hand and, adult controls using their preferred hand.

| Group | Pose | Symmetry Metrics |  |  |
| --- | --- | --- | --- | --- |
|  |  | Angle | Distance | Orientation |
| CP (Less Affected Hand) | Muscles | $0.83 \pm 0.19$ | $0.83 \pm 0.11$ | $0.91 \pm 0.06$ |
| | Powerbars | $0.85 \pm 0.15$ | $0.85 \pm 0.09$ | $0.9 \pm 0.06$ |
| CP (Affected Hand) | Muscles | $0.79 \pm 0.2$ | $0.8 \pm 0.14$ | $0.9 \pm 0.06$ |
| | Powerbars | $0.85 \pm 0.14$ | $0.87 \pm 0.1$ | $0.9 \pm 0.04$ |
| Typically developing children | Muscles | $0.9 \pm 0.1$ | $0.91 \pm 0.07$ | $0.95 \pm 0.03$ |
| | Powerbars | $0.93 \pm 0.05$ | $0.93 \pm 0.05$ | $0.95 \pm 0.04$ |
| Typically developed Adults | Muscles | $0.95 \pm 0.05$ | $0.93 \pm 0.05$ | $0.97 \pm 0.03$ |
| | Powerbars | $0.96 \pm 0.04$ | $0.95 \pm 0.03$ | $0.96 \pm 0.02$ |

### 3.1 The relationship between between angle, distance and orientation symmetries: Why all three measures are needed

In order to measure the degree to which one multi-joint arm pose is mirror symmetric to the other in 3D space, at least three independent variables are needed. Conceptually, this can be visualized by considering the poses in Figure 4. In Figure 4A, the angles subtended by the left and right arms at the shoulder and elbow joints are approximately equal. However, the two arms are clearly not mirrored poses of each other. Similarly, in Figure 4B the joint distances (distance of wrist and elbow joints from the chest or neck) between the left and right arms are approximately equal but again, the arm poses are clearly not mirrored. Also even when both, the joint angles and distances are approximately the same as shown in Figure 4C, the poses still need not be mirror symmetric. In this case the loss of symmetry is due to the fact that the positions of the left and right joints are not aligned in space (low orientation symmetry). However, it is possible to have a high orientation symmetry as shown in Figure 4D, but still have mismatched arm poses.

**Figure 4:**
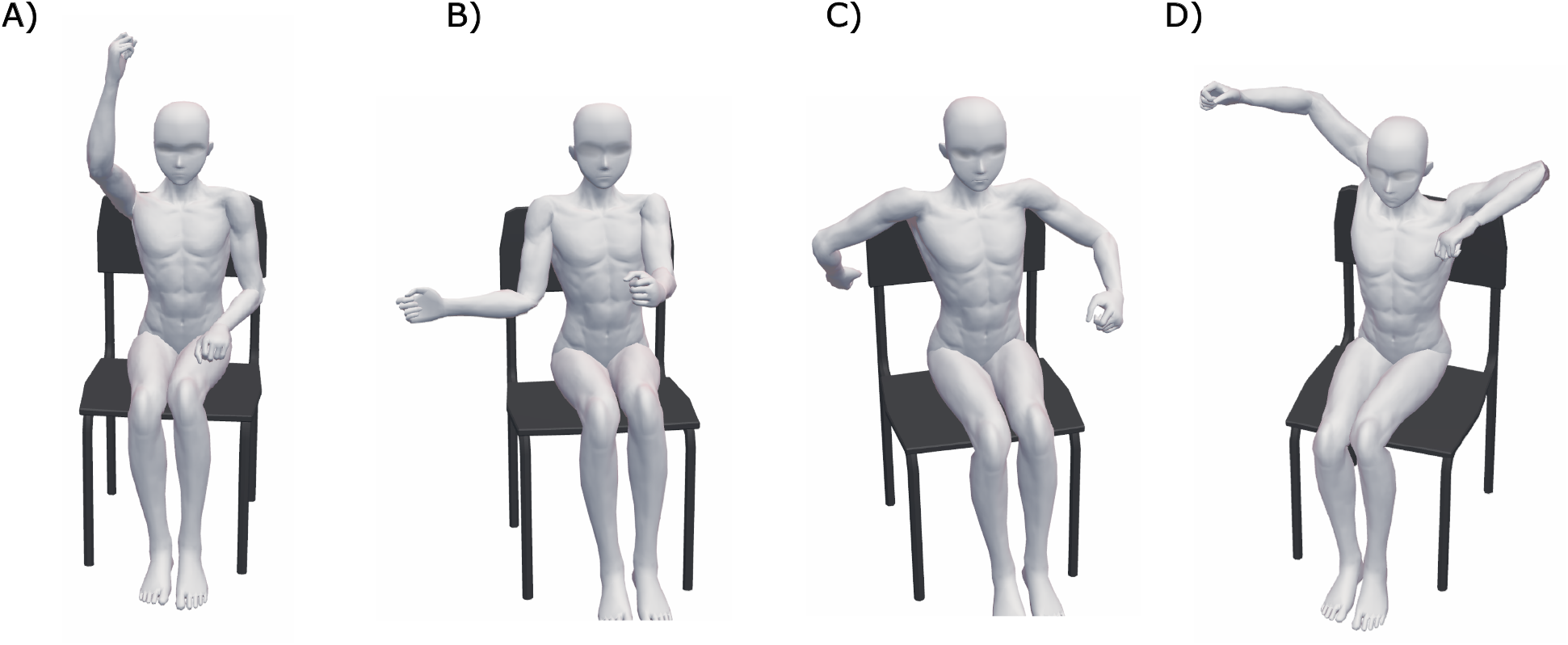
Example Poses when symmetry measures do not correlate **A.** Only distance symmetry is high (>0.9) while angle and orientation symmetries are low (< 0.3). **B**. Angle symmetry is high (>0.9) while distance and orientation symmetries are low (< 0.3). **C**. Angle and distance symmetries are high (>0.9) while orientation symmetry is low (<0.5). **D**. Orientation symmetry is high (>0.9) while angle and distance symmetries are low (<0.5)

Using a simulation of 100,000 random bilateral arm poses generated using a simple skeleton model of the shoulders, elbows, and wrists we observed weak correlations between orientation symmetry and angle symmetry (r = 0.066) as well as between orientation symmetry and distance symmetry (r = 0.284). The correlation between angle and distance symmetries were moderate with a correlation coefficient r = 0.57. These results indicate that angle and distance symmetries are complementary and semi-interdependent. However, orientation symmetry is almost independent of the other two metrics. In our simulations, we also noted that when all three metrics were high (>0.9), the resulting poses were visually mirror symmetric.

### 3.2 Assessment of multi-joint proprioceptive position sense in children with USCP

We examined the symmetry metrics as an estimate of PPS in children with USCP. Here, for each pose, the UE pose matching was done twice: first with the less affected hand then followed by the affected hand. As seen in Figure 5, we observed no significant differences in the symmetry metrics between hand poses when done with the less affected hand and the affected hand (P ≥ 0.4). The angle, distance and orientation symmetry metrics (median and standard deviation) across hands and poses are summarized in table 2.

**Figure 5:**
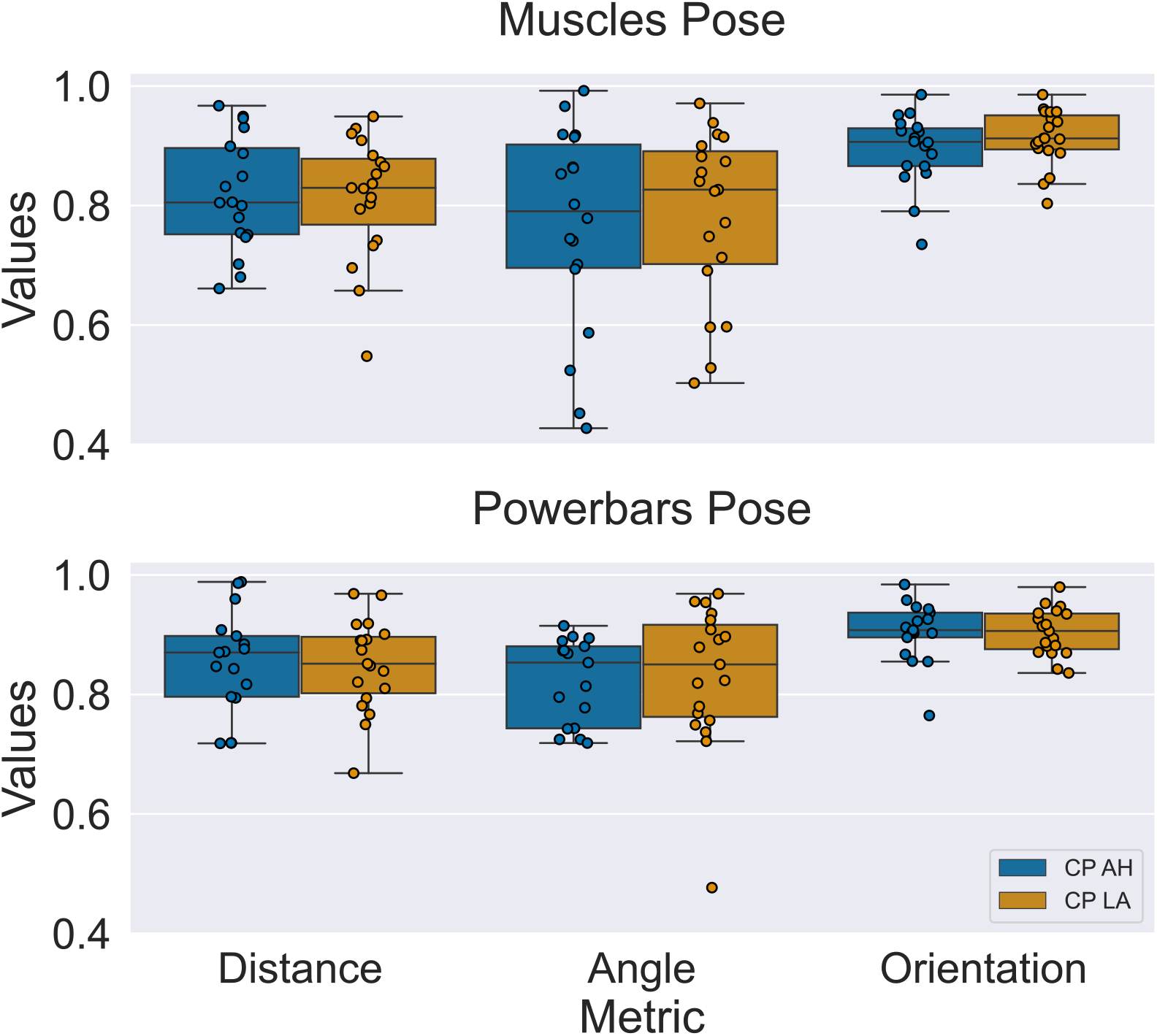
Median and interquartile ranges of symmetry metrics for children with USCP when matching the ’Muscles Pose’ (top) and ’Powerbars’ Pose (bottom) using their affected hand (CP AH) or less affected hand (CP LA). There were no statistical differences observed between the symmetry metrics when the matching was performed with the affected hand vs. the less affected hand.

We also determined if the pose type, ’Muscles’ vs ’Powerbars’, affected the measured symmetries in children with USCP. We found that the corresponding symmetries for each pose were in very similar ranges, as seen in table 2, and that there were no significant differences in corresponding symmetries between the two poses(P > 0.3).

With a subset of our participants with USCP (n=8), we also investigated if the relationship between PPS scores and the absence or presence of vision during the task. This was done to verify if the symmetry scores obtained during blindfolding were truly representative of PPS, or if there were additional neuromuscular constraints affecting the scores such as muscle fatigue or spasticity. Our symmetry score findings across 8 children with USCP performing the ’Muscles’ and ’Powerbars’ pose matching with their affected hand with and without blindfolding is detailed in Table 3. There were no statistically significant differences between the symmetry measures for blindfolded vs unblindfolded children, with P ≥ 0.38 for all comparisons.

**Table 3:** Median and interquartile ranges of Symmetry metric for PPS estimation when pose matching is done by children with USCP using their affected hand while blindfolded and when unblindfolded.No statistically siginificant differences in symmetry scores were observed between the two conditions.

|  | Pose | Symmetry Metrics |  |  |
| --- | --- | --- | --- | --- |
|  |  | Angle | Distance | Orientation |
| Blindfolded | Muscles | $0.82 \pm 0.34$ | $0.79 \pm 0.18$ | $0.93 \pm 0.03$ |
| | Powerbars | $0.85 \pm 0.1$ | $0.87 \pm 0.02$ | $0.93 \pm 0.04$ |
| Unblindfolded | Muscles | $0.71 \pm 0.18$ | $0.82 \pm 0.07$ | $0.92 \pm 0.03$ |
| | Powerbars | $0.83 \pm 0.1$ | $0.88 \pm 0.05$ | $0.93 \pm 0.03$ |

### 3.3 Assessment and comparison of multi-joint proprioceptive position sense in children with USCP in comparison to typically developing children

We next compared the symmetry metrics between children with USCP and typically developing children (TDC) during multi joint pose matching tasks. The children with USCP performed the tasks by first matching with their affected hand and then again with their less affected hand. We measured symmetry metrics from both the affected and less affected hand in children with USCP when comparing against TDC.

#### 3.3.1 PPS of children with USCP matching poses with their affected hand compared with typically developing children

When children with USCP performed the pose matching using their affected arm they again exhibited significantly lower scores for all three symmetry measures in comparison to typically developing children (Figure 6). For the ’Muscles’ pose, as seen in Table 2, the angle symmetry was observed to be higher (P=0.023) in typically developing children. Typically developing children also exhibited statistically significantly higher values of distance symmetry (P=0.025) and orientation symmetry (P=0.002). Again, for the ‘Powerbars’ pose, typically developing children had significantly higher angle (P=0.001), distance (P=0.006), and orientation (P=0.007) symmetries. These results, align with observations from previous studies that children with USCP may have lower levels of proprioceptive position sense compared to typically developing children.

**Figure 6:**
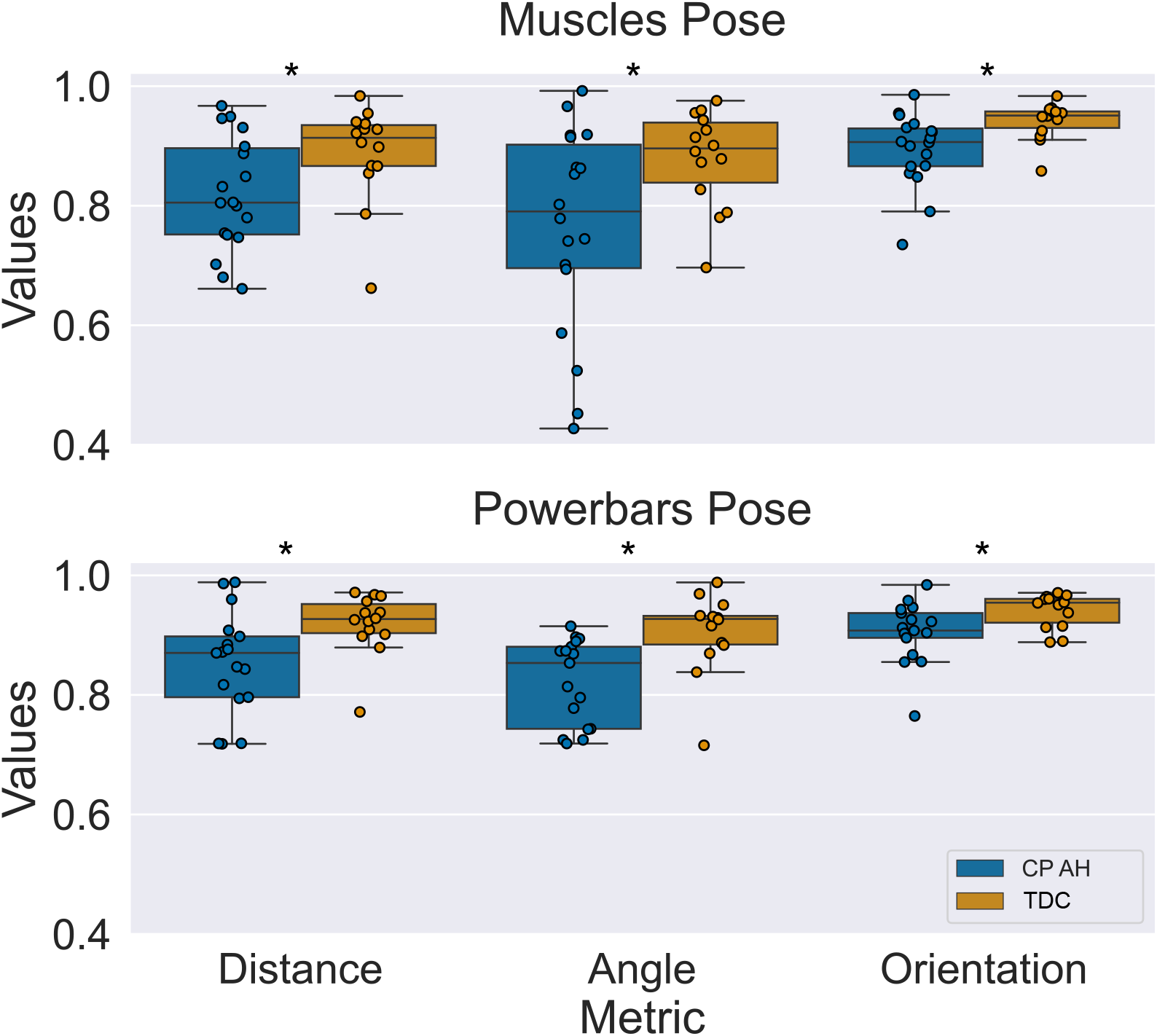
Median and interquartile ranges of PPS symmetry metrics for children with USCP performing the task with their affected hand (CP AH) as compared to age matched typically developing children (TDC). All three symmetry metrics had statistically significant differences between the CP and TDC groups for both ’Muscles’ and the ’Powerbars’ pose.

#### 3.3.2 PPS of children with USCP matching poses with their less affected hand compared with typically developing children

We again observed that when children with USCP performed the pose matching using their less affected arm, they exhibited significantly lower scores for all three symmetry measures in comparison to typically developing children,as shown in Figure 7. For the matching of the ’Muscles’ pose using the less affected hand, typically developing children had a higher angle symmetry (P=0.02) compared to children with USCP. Typically developing children also exhibited higher distance (P=0.007) and orientation (P=0.024) symmetries compared to children with USCP. In the case of matching during the ’Powerbars’ pose with the less affected arm, children with USCP again had significantly lower angle (P=0.03), distance (P=0.002), and orientation (P=0.002) symmetry scores compared to typically developing children.

**Figure 7:**
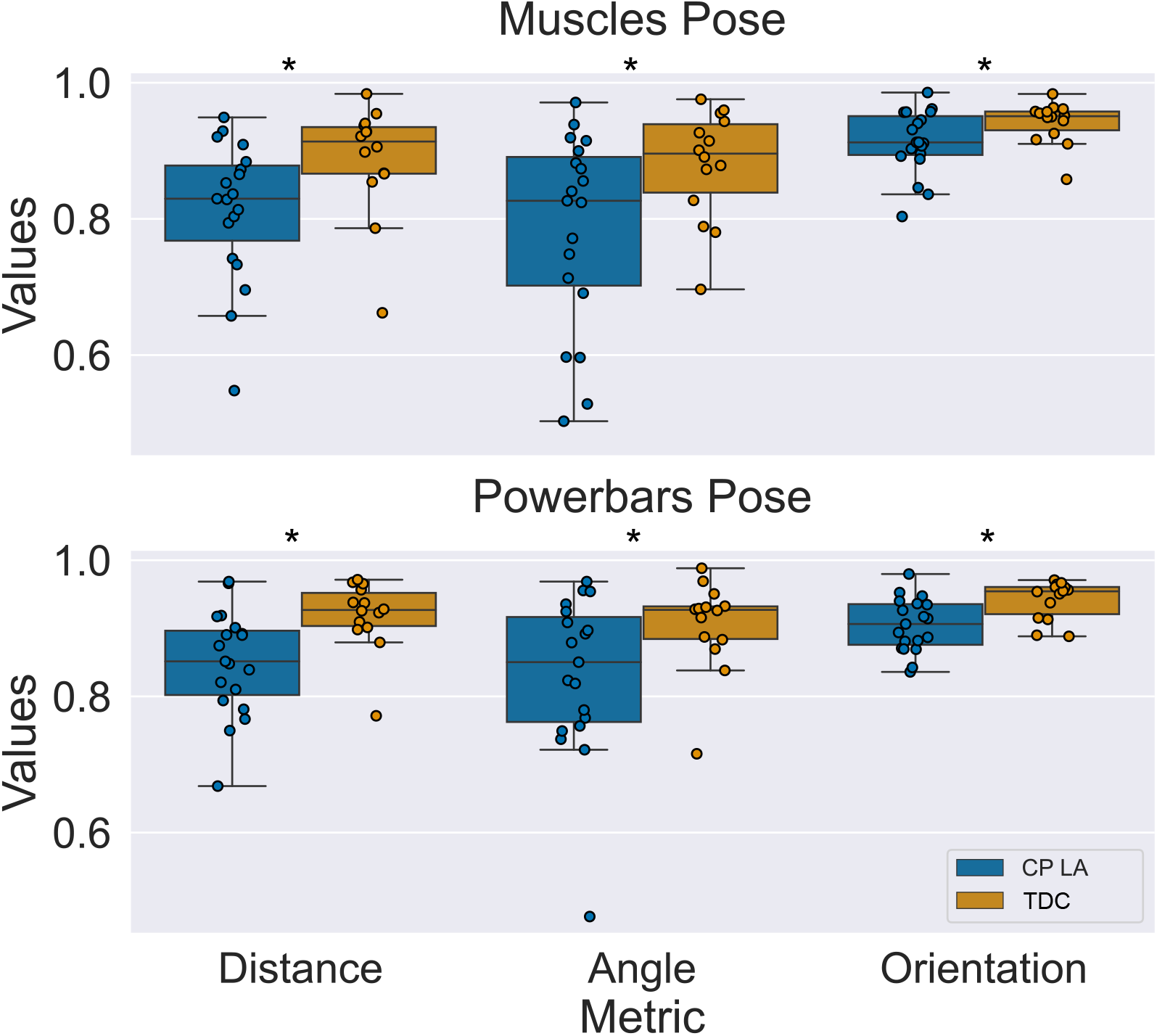
Median and interquartile ranges of PPS symmetry metrics for children with USCP performing the matching with their less affected hand (CP LA)as compared to age matched typically developing children (TDC).

### 3.4 PPS of typically developing controls compared with typically developed Adults

We performed additional analyses to determine if age was a possible factor that affects estimated PPS symmetry measures. We compared TDC aged 11 ± 3 years against typically developed adults (TDA) 32 ± 6 years for the ’Muscles’ pose and the ’Powerbars’ pose as shown in Figure 8. We observed that for the ’Muscles’ Pose, TDC had lower angle (P=0.07), distance (P=0.1) and orientation (P=0.064) symmetry measures compared to TDA. However the differences were not statistically significant. For the ’Powerbars’ pose, we observed that TDC had statistically significant lower angle symmetry (P=0.043) compared to TDA. TDC also had lower distance (P=0.13) and orientation (P=0.12) symmetries that were not statistically significant compared to TDA. These observations suggest that there is no clear trend relating age of typically developing/developed individuals and PPS symmetry metrics.

**Figure 8:**
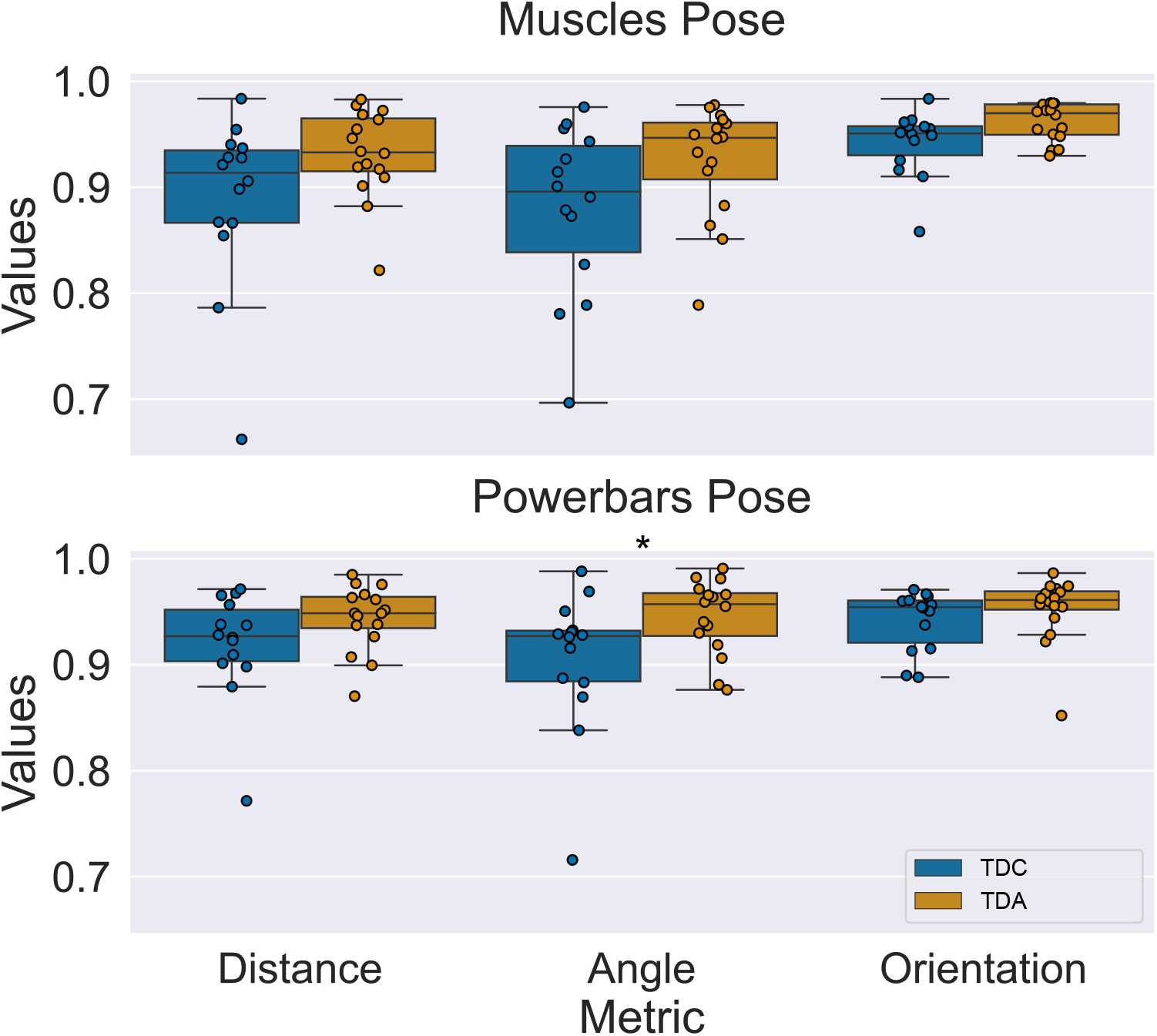
Median and interquartile ranges of PPS symmetry metrics for typically developing children compared to typically developed adult controls. While most of the symmetry metrics were not statistically significantly different between the two groups, in general adults did have slightly higher values of symmetry metrics along with less variability.

### 3.5 Relationship between PPS and hand function in children with USCP

We additionally investigated how these estimated measures of PPS correlate with hand function in children with USCP. We considered unimanual hand function assessments: the Box and Blocks Test (BBT) and the Jebson-Taylor Test of Hand Function (JTTHF) as well as bimanual hand function assessments: the Assisting Hand Assessment (AHA). Additionally, we correlated our estimates of PPS with the Manual ability Classification System (MACS) which is a questionnaire used to classify how children with cerebral palsy use their hands to handle objects in daily life.

In Figures 9 A and B, we present the absolute magnitude of correlations between symmetry measures and hand function assessment scores for the ’Muscles’ pose and the ’Powerbars’ pose, respectively. In the figures, we use (-) to denote negative correlations. In general, we see weak to moderate correlations between distance/angle symmetries and hand fuction assessment scores. However we see moderate to strong correlations between orientation symmetry and BBT, JTTHF, MACS and AHA. We observe that when the matching of the ’Muscles’ pose is performed with the affected hand, we see moderate correlations of the orientation symmetry with JTTHF assessment of the affected hand (r = -0.5), JTTHF assessment of the less affected hand (r = -0.51), and the AHA (r = 0.45). However, the orientation symmetry with the BBT is low (r ¡ 0.2) for both the affected and less affected hands. We observe that when matching is done with the less affected hand for the ’Muscles’ pose, orientation symmetry has a strong negative correlation with JTTHF assessments of both the affected (r = -0.6) as well as the less affected hand (r = -0.5). This may be due to the fact that in our approach the symmetry metrics depend on PPS abilities of both the affected and less affected hands. We also observe large positive correlations between orientation symmetry (’Muscles’ pose; less affected hand) with AHA scores (r = 0.7), MACS (r = -0.7) and BBT scores when the affected hand is used (r = 0.59).

**Figure 9:**
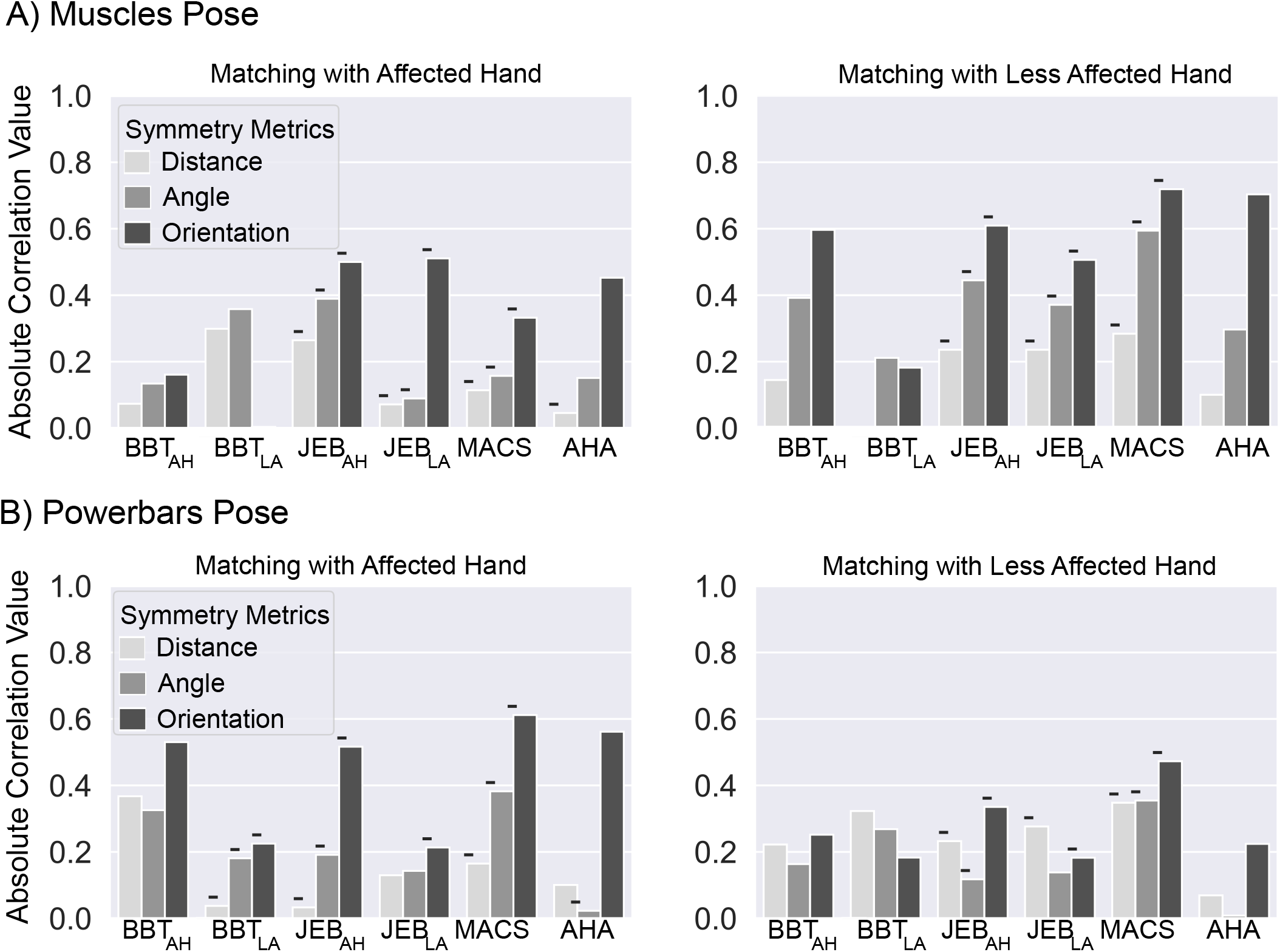
Absolute magnitude of correlation coefficients between symmetry measures and clinical assessments for the ’Muscles’ pose (A) and the and ’Powerbars’ pose (B). The left half of each figure shows the strength of correlations between the following clinical scores: Box and Blocks test (BBT), Jebsen-Taylor (JEB), Manual ability Classification System (MACS), and the Assisting Hand Assessment (AHA) against the measured symmetry metrics when the pose matching was performed with the affected hand. Similarly, the right half of each figure indicates the strength of correlations between the clinical scores and symmetry metrics when the pose matching was performed with the less affected hand. Negative correlations are indicated with the (-) symbols adjacent to the respective bars. All correlations were calculated using Spearman’s rank correlation.

In the case of the ’Powerbars’ pose, when matching with the affected hand, orientation symmetry correlated moderately with the JTTHF scores with the affected hand (r = -0.51) and BBT scores with the affected hand (r=0.53). The orientation symmetry had even stronger correlations with MACS (r=-0.61), and the AHA (r=0.56). In contrast, when the matching for the ’Powerbars’ pose was done using the less affected hand, we observed that the orientation symmetry had much weaker correlations overall, with the only moderate one being with MACS (r = -0.47).

## 4 Discussion

Estimating overall upperlimb proprioceptive position sense (PPS) in an individual, requires the consideration of multiple-joints that can move without constraint in 3D space as well as a comprehensive system of measurements. Using a large number of simulated random arm poses we showed in section 3.1 that for an upperlimb contralateral pose matching task in 3D space, three distinct measurements: angle, distance and orientation symmetry had to be considered together to completely describe the similarity or mirroring ability (as an estimate of PPS) of an individual’s pose. Applying these measures to participants, we consistently observed that all three symmetry measures were lower in children with USCP compared to TD children (Figures 6 and 7). We also found that while angle and distance symmetry were not particularly indicative of hand function abilities, orientation symmetry exhibited strong correlations with clinical hand function assessments scores in children with USCP as shown in Figure 9.

Previous studies have reported that proprioception, as estimated and measured by various methods, is compromised in individuals with hemiplegic cerebral palsy [8, 30, 31]. In our study we confirmed this observation that children with USCP exhibit estimated PPS deficits compared to typically developing age matched controls. Additionally, we consistently observed that the group of children with USCP exhibit increased variabilities in all three symmetry measures compared to typically developing age matched controls. Similar observations of increased variability in estimated proprioception across children with cerebral palsy were made in previous studies where PPS was estimated using a robotic device [9], or movement threshold detection [32]. These findings suggest that while children with cerebral palsy tend to exhibit comparatively lower values of PPS compared to typically developing children, the degree to which PPS is compromised may vary significantly from child to child. We also observed that angle and distance symmetry were generally weakly correlated with scores from unimanual tests: the box and blocks test (BBT) and the Jesbsen Taylor Test for Hand Function (JTTHF) with |r| < 0.4. We similarly observed that the bimanual hand function, Assisting Hand Assessment (AHA), scores weakly to moderately correlated with angle and distance measures of multi-joint PPS with r < 0.4. These observations are in line with those from a previous study by Arnould et al., where the correlation between estimated proprioception and BBT was weak, with r = 0.24 for the less affected hand and r = 0.36 for the more affected hand [7]. Other studies [33] have similarly reported weak to moderate correlations between assessed proprioception and hand function as measured by the AHA (r = 0.4) and MACS (r = 0.49).

However, there have been reports of the opposite finding from studies that have used kinematic information via robotic devices during active tasks have reported moderate to strong correlations between certain proprioceptive measurements and hand function assessments [9]. In our study too, we found that the orientation symmetry measure alone was consistently strongly correlated with hand function ability for both unimanual (BBT, JTTHF) and bimanual (AHA) assessments.

Contradictory findings from a number of studies have led researchers to conclude that the relationship between PPS and hand function is still poorly understood [5]. Based on observations from our study we suggest that these contradictory findings may partly arise due to differences in PPS measurement methods and specifically how PPS is affected in children with USCP. We argue that single metrics of PPS measurements from a single joint may not necessarily completely represent the PPS ability of an individual. This observation is inline with more recent reports of proprioception being a high level sensorimotor system rather than a purely sensory system [13]. Despite the fact that children with USCP exhibit relatively lower values of estimated PPS via all three of our symmetry measures, it is orientation symmetry alone that shows trends of correlation with hand function. Upper limb orientation symmetry, as introduced in Section 2.4.3, measures how well coordinated multiple joints on both sides are. Hand function abilities are also highly dependent on coordination between different joints within and across each side of the body. Additionally, in order for the body to exhibit greater orientation symmetry, sensory and motor information of multiple joints from both the reference and matching arms need to be reliably integrated. Considering this along with our findings, we suggest that in children with USCP, not only is information from different sensory receptors compromised, but the ability to **integrate** multiple sources of positional information to predict position may also be affected.

Although our study provides methods for estimate PPS in children with USCP and insights into the relationship between PPS and hand function, there are some of limitations in the study. Our approach to estimating multi-joint upperlimb PPS in individuals relies on unconstrained and unassisted limb movements. This approach may not be pragmatic for assessing PPS in those who have difficulties moving their limbs or in suspending their arms stationarily for a few seconds. In such cases, it may still be possible to estimate PPS using our methods if assistive but unconstrained arm support via exoskeletons can be used. Another limitation in this study is the fact that we only use a single trial for each pose matching task. While, this enables quicker estimation while minimizing fatigue and boredom in young children, the reliability of intra-subject PPS measurements still needs to be determined to identify the minimum number of trials per task needed to accurately estimate PPS. Additionally, while the measurement of PPS using our approach may be quicker and more comprehensive than traditional methods, this study was done using kinematic data acquired from a sophesticated opto-electronics system that required reflective markers to be be expertly placed on bony landmarks. Due to the size and cost, access to such equipment may be limited to some researchers. We plan to address this in future work by using the same PPS measurement metrics but with kinematic data acquired from low cost marker-less video based motion capture systems [34].

In this study, we proposed an effective way to estimate multi-joint upper limb proprioceptive position sense in individuals by using kinematic data captured during an unconstrained contralateral joint pose matching task in three-dimensional space. We found that at least three different metrics for measuring pose similarity were needed to quantify multi-joint pose matching in 3D space. We also showed that our proposed method of estimating proprioceptive position sense could detect group level differences between children with unilateral spastic cerebral palsy and typically developing children. Additionally, we observed that coordination of joints during the pose matching task which was represented by orientation symmetry strongly correlated with hand function assessment scores in children with unilateral spastic cerebral palsy. We suggest that angle and distance symmetries may be affected by the reliability of low-level sensory information from mechanoreceptors and proprioceptors while orientation symmetry represents a high-level assimilation of sensory and motor information that is critical for hand coordination.

## Data Availability

All data produced in the present study are available upon reasonable request to the authors.

## 5 Acknowledgements

This research was supported by the Eunice Kennedy Shriver National Institute Of Child Health & Human Development of the National Institutes of Health under Award Numbers R01HD095663 and R01HD076436.

